# Dynamic change and clinical relevance of post-infectious SARS-CoV-2 antibody responses

**DOI:** 10.1101/2021.01.24.20248381

**Authors:** PWG Mallon, W Tinago, A Garcia Leon, K McCann, G Kenny, P McGettrick, S Green, R Inzitari, AG Cotter, ER Feeney, S Savinelli, P Doran, on behalf of the All Ireland Infectious Diseases Cohort Study Group.

## Abstract

**Background:** Although reports suggest that most individuals with COVID-19 develop detectable antibodies post infection, the kinetics, durability, and relative differences between IgM and IgG responses beyond the first few weeks after symptom onset remain poorly understood.

**Methods:** Within a large, well-phenotyped, diverse, prospective cohort of subjects with and without SARS-CoV-2 PCR-confirmed infection and historical controls derived from cohorts with high prevalence of viral coinfections and samples taken during prior flu seasons, we measured SARS-CoV-2 serological responses (both IgG and IgM) using commercially available assays. We calculated sensitivity and specificity, relationship with disease severity and mapped the kinetics of antibody responses over time using generalised additive models.

**Results:** We analysed 1,001 samples from 752 subjects, 327 with confirmed SARS-CoV-2 (29.7% with severe disease) spanning a period of 90 days from symptom onset. Sensitivity was lower (44.1-47.1%) early (<10 days) after symptom onset but increased to >80% after 10 days. IgM positivity increased earlier than IgG-targeted assays but positivity peaked between day 32 and 38 post onset of symptoms and declined thereafter, a dynamic that was confirmed when antibody levels were analysed, with more rapid decline observed with IgM. Early (<10 days) IgM but not IgG levels were significantly higher in those who subsequently developed severe disease (signal / cut-off 4.20 (0.75-17.93) versus 1.07 (0.21-5.46), P=0.048).

**Conclusions:** This study suggests that post-infectious antibody responses in those with confirmed COVID-19 begin to decline relatively early post infection and suggests a potential role for higher IgM levels early in infection predicting subsequent disease severity.

## Introduction

Severe acute respiratory syndrome coronavirus 2 (SARS-CoV-2), the cause of COVID-19 was first identified in December 2019 in Wuhan, China before rapidly becoming pandemic. Over and above the significant proportion of asymptomatic cases, the majority of symptomatic COVID-19 cases are mild. However, up to 20% of infections progress to severe disease, as classified by the World Health Organisation (WHO),[1] with comorbidities, male sex and older age associated with poorer outcomes.[2] It remains unclear to what extent infection with SARS-CoV-2 confers post-infectious immunity, either through humoral (antibody-mediated) or cellular (T-cell mediated) mechanisms.

Emerging data suggest that many individuals, particularly with severe COVID-19, mount detectable anti-SARS-CoV-2 IgG responses within two weeks after infection,[3] with many factors influencing antibody responses including age, disease severity and time from onset of symptoms, with variable intensity and durability of serologic responses reported.[4, 5] Serology plays important roles in the diagnosis of many infections, both at an individual and population level, with early IgM responses used to detect recent infections and more persistent memory IgG responses used to estimate seroprevalence. However, given the uncertainties surrounding development and persistence of antibody responses to SARS-CoV-2,[6] the role of serology in diagnosis or surveillance of COVID-19 remains to be fully clarified.

A number of commercial anti-SARS-CoV-2 serological assays report high sensitivity and specificity. However, their validation in real-world settings, taking into account the range of factors that affect serologic responses, including cross-reactivity against other chronic infections, has been limited.[7]

Although several studies have compared commercially available serological assays in COVID-19, many have small sample sizes[8] or lack non-SARS-CoV-2 infected controls.[7, 9] Additionally, inclusion of uninfected controls defined as not detected on SARS-CoV-2 PCR [10] raises the potential for false positive antibody tests to be misinterpreted in those with previous infection, particularly where detailed clinical information is lacking. Many other studies either have limited data on disease severity[11, 12], or have over-representation of hospitalized patients with severe disease. In one systematic review only 4 of 40 studies included non-hospitalised patients, [13] which limits the generalisability of some observations, such as associations between higher antibody titres and disease severity.[14] In one of the largest studies to date, analysing 976 pre-pandemic blood samples and 536 blood samples from patients with SARS-CoV-2 infection, severity data was only available for 29%.[15] Lastly, although SARS-CoV-2 serological responses are dynamic, not all studies either report or account for time since symptom onset; in a recent Cochrane systematic review, 19 of 57 included studies did not stratify by time since symptom onset.[16] The same review found very little data beyond 35 days post onset of symptoms.

To address these data gaps, we aimed to compare several different commercial SARS-CoV-2 serological assays in demographically, clinically diverse and well phenotyped clinical cohorts, to define the dynamic change in qualitative and quantitative antibody responses over time since symptom onset, and delineate the relative role of IgM versus IgG antibodies in relation to onset and severity of infection in clinical samples from individuals with and without COVID-19 infection.

## Methods

### Study design

The All Ireland Infectious Diseases (AIID) Cohort study is a prospective, multicentre cohort enrolling patients attending clinical services for Infectious Diseases in Ireland. Subjects provide data on demographics (age, sex, ethnicity, smoking status), clinical characteristics (hospitalisation, date of symptoms onset, underlying conditions e.g. diabetes, malignancies) and laboratory results. COVID-19 disease severity was defined according to the WHO guidance[1] and collapsed into severe and non-severe for analysis. Subjects also provided blood samples for bio-banking on up to three occasions. All subjects provide written, informed consent and the study is approved by local and national research ethics committees.

For this analysis we included AIID Cohort participants who presented to the Mater Misericordiae University Hospital and St Vincent’s University Hospital with symptoms consistent with COVID-19 between the 26th of March 2020 and the 10th of July 2020 with available biobanked samples. In addition, as controls, we included subjects with plasma samples biobanked prior to 2020 from the AIID Cohort and another longitudinal study of subjects with and without HIV infection,[17] including samples specifically taken during previous flu seasons from 2016 to 2019, as outlined in national statistics.[18, 19]

### Laboratory analysis

Plasma, stored at −80°C and thawed in batches, underwent same-day serological testing in the Core Laboratory in the Clinical Research Centre, University College Dublin, Ireland by blinded technicians using four assays according to manufacturers’ instructions: the Elecsys® anti-SARS-CoV-2 electrochemiluminescence immunoassay (Roche Diagnostics, Penzberg, Germany) run on the Cobas® e411 automated platform (Roche Diagnostics), the SARS-CoV-2 IgG chemiluminescent microparticle immunoassay (CMIA) (Abbott Laboratories, IL, USA) run on both the Architect i2000SR platform (Abbott Diagnostics), the Abbott Alinity ci platform (Abbott Diagnostics) and the Abbott SARS-CoV-2 IgM assay run on the Abbott Architect i2000SR platform. The Elecsys assay is a sandwich immunoassay that detects IgA, IgM and IgG antibodies to SARS-CoV-2, so a positive result may reflect reactive, non-IgG antibody responses. The Abbott SARS-CoV-2 IgG and IgM assays are two-step immunoassays targeting the SARS-CoV-2 nucleocapsid protein.

The Elecsys assay presents results as a cut-off index (COI), derived from comparison of electrochemiluminescence signals from the sample to positive and negative calibration signals with both qualitative (reactive or non-reactive) and quantitative (COI) results provided. The Abbott assays also provide automated qualitative and quantitative (signal/cut-off (S/CO) ratios) results. For the IgG and IgM assays, S/CO ≥1.4 and ≥1.0 respectively were interpreted as reactive. The Alinity and Architect versions of the Abbott IgG assay are similar; both use the same capture antibody, conjugate material and the same formulations of calibrator and QC material, with corresponding ranges.

### Statistical Analysis

Continuous and categorical variables are summarised using median with interquartile range (IQR) and frequency/percentage respectively. Sensitivity and specificity along with their binomial exact 95% confidence intervals (CI) were used to describe the performance characteristics of the assays. Sensitivity was calculated based on samples from subjects who tested detected on SARS-CoV-2 PCR (SARS-CoV-2 Pos). Specificity was derived for two distinct groups (i) samples from subjects who presented for hospital care during the 2020 pandemic but who tested ‘not detected’ on SARS-CoV-2 PCR (SARS-CoV-2 Neg) and (ii) historical controls-subjects (Controls Pre-2020) that included subjects with and without chronic infections such as HIV and hepatitis C as well as subjects with biobanked serum samples taken during prior flu seasons between 2016 and 2019.

Within the SARS-CoV-2 Pos group, assay sensitivity was also evaluated at different time periods after date of symptom onset; 0-10, 11-21, 21-42 and >42 days. We used scatter plots with superimposed curves fitted using generalised additive models (GAM), with either a Gaussian or binomial link function and time since symptom onset fitted as a spline, to depict the non-linear relationship between time from symptom onset with either (i) quantitative antibody levels or (ii) seropositivity rate as dependent variables respectively. We compared quantitative antibody responses (COI for Elecsys or S/CO for Abbott IgG and IgM assays, referred to as antibody ‘levels’) and positivity rate for the first two time periods post symptoms onset (0-10 and 11-21 days) between subjects categorised into severe and non-severe maximal disease stage attained using the Wilcoxon rank sum test and the chi-square test respectively.

Overall sensitivity and specificity were compared between assays using the McNemar’s chi-square test as previously described.[20, 21] Overall concordance between the assays was evaluated using the Cohen’s Kappa and percentage agreement. Cross-reactivity was assessed in the Controls Pre-2020 group in samples from subjects with and without known chronic viral infections (HIV, hepatitis C or B) and samples from the 2016-2019 flu seasons. All analyses were conducted using Stata 15 (College Station, Tx) and R version 4.0.2.

## Results

A total of 752 subjects provided 1,001 samples for analysis. The SARS-CoV-2 Pos group comprised 202 individuals who provided 327 samples between March 26^th^ and July 10^th^ 2020, the SARS-CoV-2 Neg group included 149 subjects who provided 222 samples. Among these two groups, 76 (37.6%) and 49 (32.9%) provided ≥2 samples respectively. Samples were collected a median (IQR) 19 (11-41) and 8 (5-17) days post symptom onset in the SARS-CoV-2 Pos and Neg groups respectively. The Controls pre-2020 group comprised 401 subjects who provided 452 samples collected prior to 2020, including 116 samples taken during previous flu seasons. Within the Controls pre-2020 group, 19 (4.8%) were hepatitis B surface antigen positive and the majority (80%) were HIV antibody positive, of whom 40 (12.5%) were also hepatitis C antibody positive (table 1).

**Table 1:** Characteristics of the study population.

|  | SARS-CoV-2<br>Pos | SARS-CoV-2 Neg | Controls<br>Pre-2020 |
| --- | --- | --- | --- |
| Characteristic | (n=202) | (n=149) | (n=401) |
| Age (years) | 57 (45-68) | 60 (42-75) | 45 (40-53) |
| Gender: Male n (%) | 108 (54.0%) | 87 (58.4%) | 234 (58.4%) |
| Ethnicity: n (%) |  |  |  |
| Caucasian | 135 (66.8%) | 126 (84.5%) | 217 (54.1%) |
| Asian | 21 (10.4%) | 4 (2.7%) | 7 (1.8%) |
| African | 2 (1.0%) | 1 (0.7%) | 143 (35.7%) |
| Unknown | 44 (21.8%) | 18 (12.1%) | 34 (8.4%) |
| Current smoker n (%) | 12 (5.9%) | 20 (13.4%) | - |
| Diabetes: Yes n (%) | 26 (12.9%) | 15 (10.1%) | - |
| Underlying malignancies n (%) | 17 (8.4%) | 4 (2.7%) | - |
| Hospital admission: Yes n (%) | 145 (71.8%) | 119 (79.9%) | - |
| COVID-19 disease severity n (%)* |  |  |  |
| Mild | 117 (57.9%) | - | - |
| Moderate | 25 (12.4%) | - | - |
| Severe | 60 (29.7%) | - | - |
| Coinfections n (%) |  |  |  |
| HIV mono infected | - | - | 270 (67.5%) |
| HIV/HCV | - | - | 40 (10.0%) |
| HIV/HBV | - | - | 11 (2.8%) |
| HBV |  |  | 8 (2.0%) |
| **CRP (mg/L) | 23.9 (1.7-76) | 27.8 (4-89) | - |
| §Ferritin (ug/L) | 376 (141-1085) | 205 (83-447) | - |

Data are median (IQR) unless otherwise stated. HBV status defined as sAg positive. HIV; Human immunodeficiency virus. HCV; hepatitis C virus. HBV; hepatitis B virus. CRP; C-reactive protein. WHO; World Health Organisation. N; number.
\*Disease severity assigned according to WHO criteria (1).
\*\*CRP missing in 4 and 15 SARS-CoV-2 Pos and SARS-CoV-2 Neg respectively.
§Ferritin missing in 7 and 18 SARS-CoV-2 Pos and SARS-CoV-2 Neg respectively.

The median (IQR) age of the SARS-CoV-2 Pos and SARS-CoV-2 Neg groups were similar (57 (45-68) years and 60 (42-75) years respectively), while the Controls Pre-2020 group were younger (45 (40-53) years). Among all three study groups, males and those of Caucasian ethnicity were most represented (Table 1). Compared to the SARS-CoV-2 Neg group, the SARS-CoV-2 Pos group were more likely to be diabetic, have an underlying malignancy and less likely to smoke. Although the majority of the SARS-CoV-2 Pos and SARS-CoV-2 Neg groups were admitted to hospital (71.8% and 79.9% respectively), within the SARS-CoV-2 Pos group, most (58%) experienced only mild disease, 12% moderate and 30% severe disease severity respectively.

### Sensitivity and specificity

Overall, the sensitivity for all four assays was relatively low, ranging from 74.3% to 77.1% (Table 2), with no significant difference in sensitivity between assays (Supplementary table 1). In contrast, all three IgG-targeted assays and the IgM assay were highly specific, ranging from 92.7% to 100% (Table 2), with the Elecsys assay (100% 95% C.I. 99.2, 100) having marginally, but significantly higher specificity than the Abbott IgG assays (IgG Architect 99.1 (97.7, 99.8), IgG Alinity 99.1% (97.7, 99.8), % difference +0.0009 (95% C.I. 0.0002, 0.018), P=0.046). Specificity did not differ between the Abbott IgG assays (Architect 99.1% (97.7, 99.8), Alinity 99.1% (97.7, 99.8), % difference +0.00 (95% C.I. −0.006, 0.006)). The percentage of agreement between the three IgG-targeting assays ranged between 96.8% and 99.7% (Supplementary Table 2).

**Table 2:** Overall sensitivity and specificity of the three immunoassays to detect antibodies against SAR-CoV-2.

| Number of serum samples |  |  |  |  |  |  |  |  |  |
| --- | --- | --- | --- | --- | --- | --- | --- | --- | --- |
| Type of assays | Sensitivity |  |  | Specificity |  |  |  |  |  |
|  | SARS-CoV-2 Pos | Antibody positive | % (95% CI) | SARS-CoV-2 Neg | Antibody negative | % (95% CI) | Controls Pre-2020* | Antibody negative | % (95% CI) |
| Elecsys | 327 | 252 | 77.1<br>(72.1; 81.5) | 222 | 213 | 95.9<br>(92.4; 98.1) | 450 | 450 | 100<br>(99.2; 100) |
| Alinity-IgG | 327 | 244 | 74.6<br>(69.5; 79.2) | 222 | 214 | 96.4<br>(93.0; 98.4) | 450 | 446 | 99.1<br>(97.7; 99.8) |
| Architect-IgG | 327 | 243 | 74.3<br>(69.2; 79.0) | 222 | 214 | 96.4<br>(93.0; 98.4) | 450 | 446 | 99.1<br>(97.7; 99.8) |
| *Architect-IgM | 323 | 247 | 76.5<br>(71.5; 81.0) | 222 | 206 | 92.7<br>(88.6; 95.8) | 116 | 115 | 99.1<br>(95.3; 100) |

When measured in all available samples, sensitivity for all assays was lower than specificity. SAR-CoV-2; severe acute respiratory syndrome coronavirus 19. Ab; antibody. CI, confidence interval. \* IgM was only measured in the control samples from prior flu-seasons (n=116) and data were available for 150 of 152 samples for each of the other three assays.

The sensitivity of the three IgG-targeted assays increased considerably with time after onset of COVID-19 symptoms; 44.1% to 47.1% in samples collected ≤10 days post symptom onset increasing to a maximum sensitivity ranging from 86.5% to 90.5% in samples collected at >42 days post symptom onset, with no significant differences between the three assay (Figure 1, Table 3). In contrast, the Abbott IgM assay sensitivity increased from a low of 57.6% in the early (≤10 days) period to a high of 89.0% at 11-21 days post symptom onset, but notbaly decline to 68.5% >42 days post symptom onset (Figure 1).

**Table 3:** Performance of the three serological assays in different periods from date of onset of symptoms.

| Days from symptom onset | Elecsys |  | Alinity-IgG |  | Architect-IgG |  | Architect-IgM |  |
| --- | --- | --- | --- | --- | --- | --- | --- | --- |
|  | Ab positive | Sensitivity (95% CI) | Ab positive | Sensitivity (95% CI) | Ab positive | Sensitivity (95% CI) | Ab positive | Sensitivity (95% CI) |
| 0-10 | 32/68 | 47.1 (34.8; 59.6) | 31/68 | 45.6 (33.5; 58.1) | 30/68 | 44.1 (32.1; 56.7) | 38/66 | 57.6 (44.8; 69.7) |
| 11-21 | 91/110 | 82.7 (74.3; 89.3) | 89/110 | 80.9 (72.3; 87.8) | 89/110 | 80.9 (72.3; 87.8) | 97/109 | 89.0 (81.6; 94.2) |
| 22-42 | 62/75 | 82.7 (72.2; 90.4) | 60/75 | 80.0 (69.2; 88.4) | 60/75 | 80.0 (69.2; 88.4) | 62/75 | 82.7 (72.2; 90.4) |
| >42 | 67/74 | 90.5 (81.5; 96.1) | 64/74 | 86.5 (76.5; 93.3) | 64/74 | 86.5 (76.5; 93.3) | 50/73 | 68.5 (56.6; 78.9) |

The proportion of individuals with SARS-CoV-2 PCR-detected diagnosis of COVID-19 with positive antibody responses was higher after 10 days from symptom onset for all assays. Ab; antibody. CI, confidence interval.

**Figure 1.**
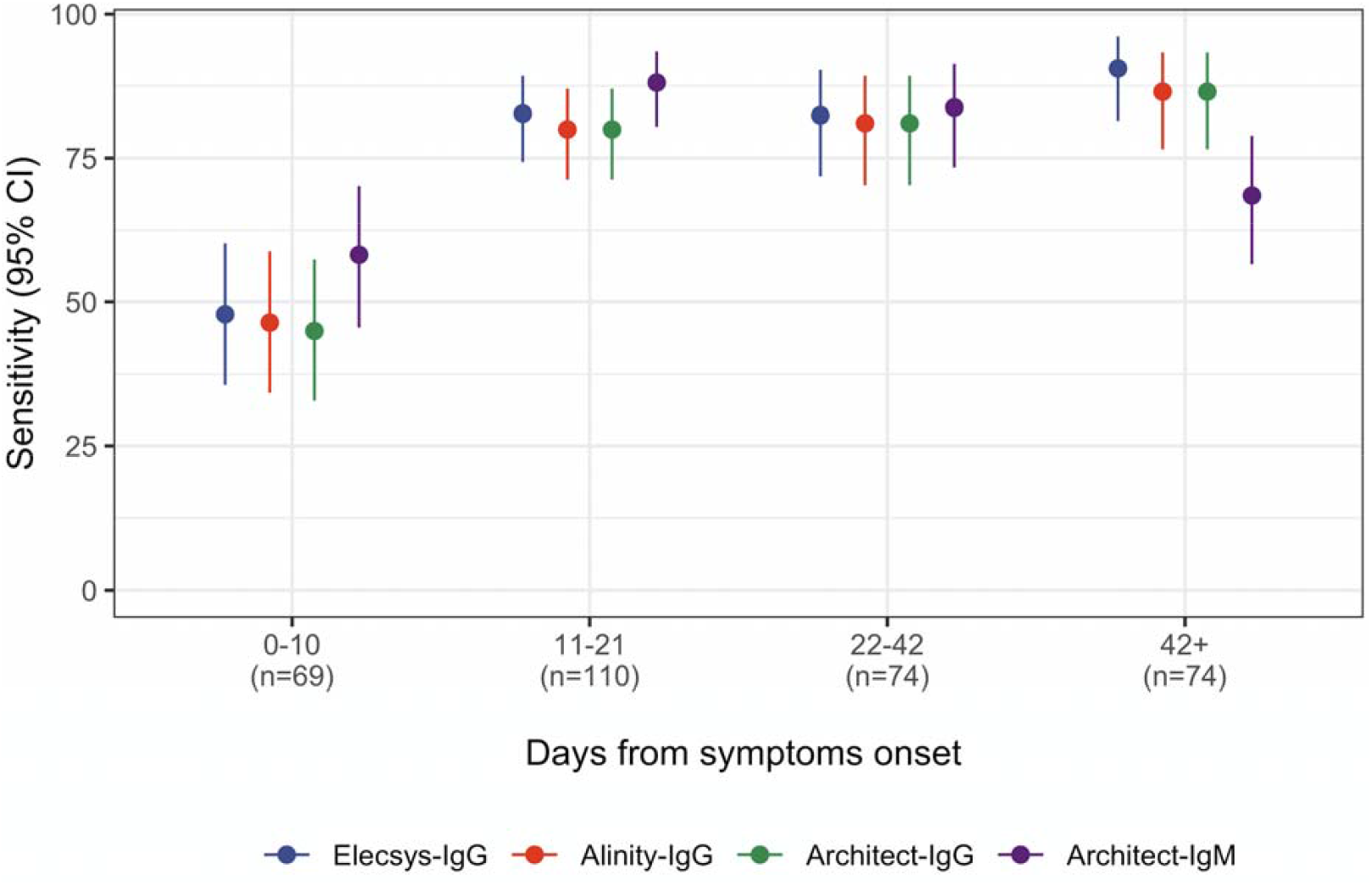
Sensitivity of serological assays based on time periods from onset of symptoms. N; number. CI; confidence interval.

Merging Abbott Architect IgM and IgG positive responses into a single variable did not appreciably alter overall sensitivity (82.9% (95% C.I. 78.3, 86.8)) or specificity (98.3% (93.9, 99.8)) or at any of the timepoints analysed (data not shown).

From fitted GEE longitudinal curves, estimated positivity rates peaked at day 38 post symptom onset for the Elecsys assay (positivity rate 92.2% (95% CI 85.2; 96.0)), day 36 for the Abbott Alinity IgG assay (positivity rate 89.1% (95% CI 81.6; 93.8)) and day 36 for the Abbott Architect IgG assay (positivity rate 89.1% (95% CI 81.6; 93.8), Supplementary Figure 1). There was a more rapid increase in the positivity rate with Abbott Architect IgM assay, with an earlier peak at day 23 post symptom onset (positivity rate 88.7% (95% CI 82.8; 92.8) Supplementary Figure 2). In addition, after day 23 from symptom onset, there was a more rapid decline in positivity rate with the Abbott IgM assay compared to the IgG assay (Figure 2).

**Figure 2.**
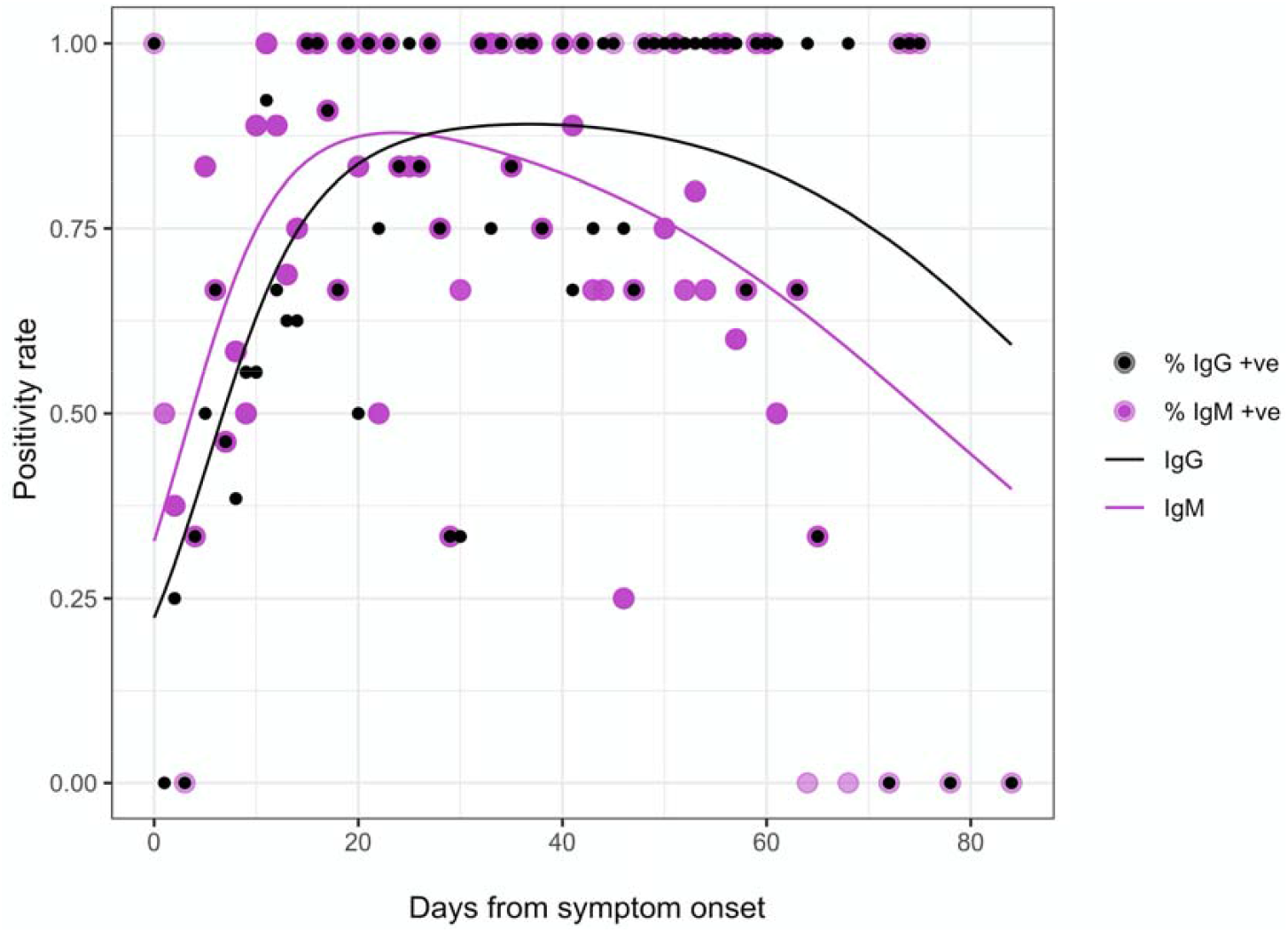
Dynamic change in seropositivity over time since onset of symptoms. Fitted probability of positive IgG/IgM antibody test estimated using GEE with logit link and time as natural spline. IgM positivity rates increase more rapidly but fall off earlier than IgG-directed serological assays.

### Dynamics of antibody levels in the SARS-CoV-2 Pos Group

Overall, median (IQR) antibody levels for the three IgG-targeted assays were 11.93 (1.66-35.64) for Elecsys, 6.47 (1.35-10.43) for Abbott Alinity and 4.71 (1.17-6.53) for Architect and for the Abbott Architect IgM was 6.35 (1.17-15.88). The Abbott Alinity and Architect assay levels were highly correlated (repeated measures correlation coefficient r=0.99 (95% CI 0.98; 0.99)), but less strongly between Elecsys and Architect (r=0.60 (0.47; 0.70) and Ececsys and Alinity (r=0.62 (0.50;0.72) respectively.

IgG-targeted antibody levels increased after onset of symptoms, peaking at 47, 35 days and 36 days post symptoms onset for the Elecsys (peak COI of 32.9), Abbott Alinity (peak S/CO 8.2) and Abbott Architect (peak S/CO 5.4) respectively (Figure 3a-c). The IgM antibody titres peaked earlier at 26 days and followed a more rapid decline thereafter relative to the other three assays (Figure 3d).

**Figure 3.**
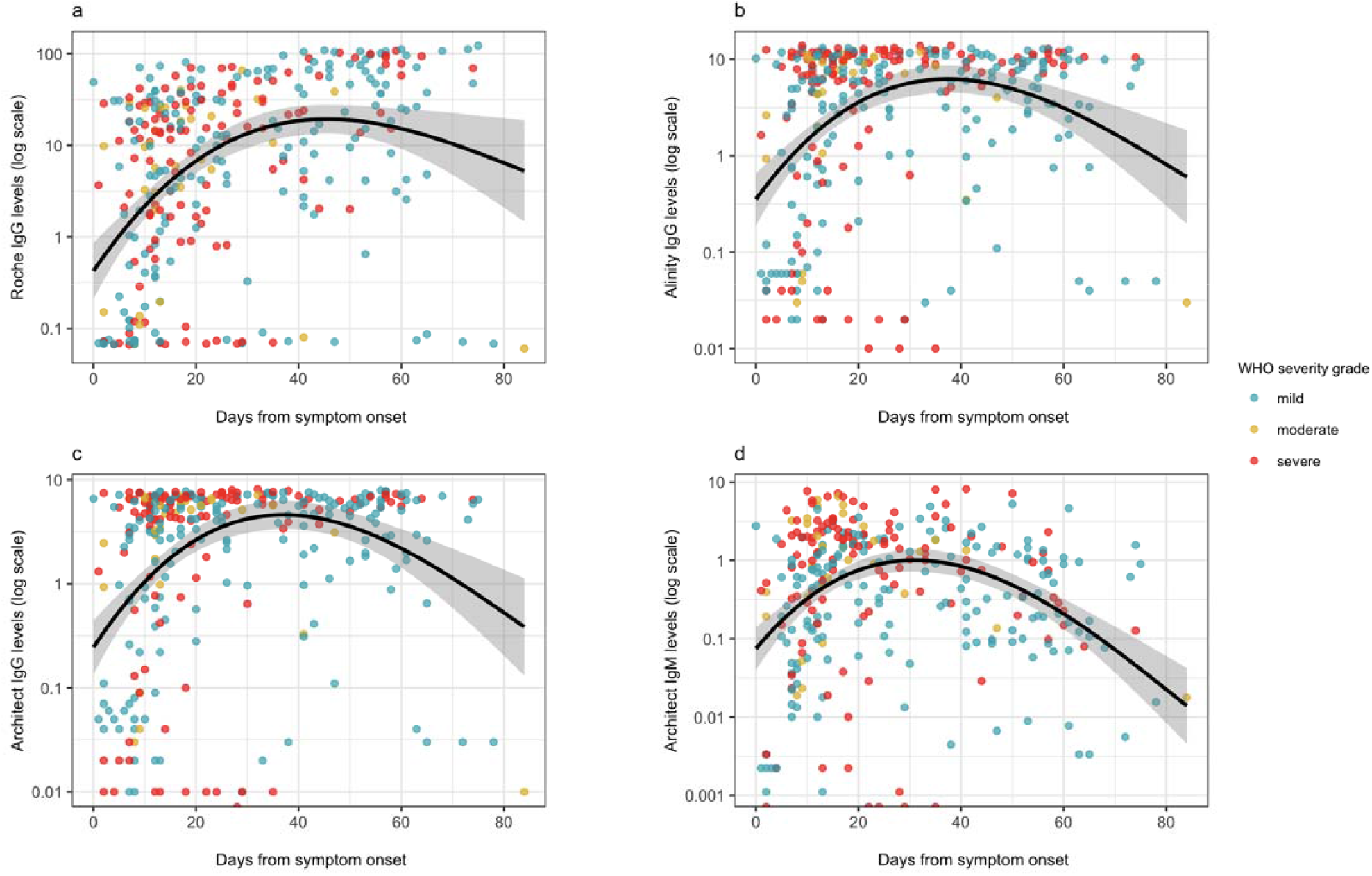
Dynamic change in antibody levels over time since onset of symptoms. Log; logarithmic

There were no significant differences in early IgG-targeted antibody levels (0-10 or 11-22 days post symptom onset) between subjects who developed severe versus non-severe COVID-19 infection (Figure 4, Supplementary table 2). In contrast, early IgM antibody levels were significantly higher in subjects who developed severe COVID-19, with an almost four-fold higher IgM at day 0-10 post symptom onset (4.20 (0.75-17.93) versus 1.07 (0.21-5.46), P=0.048). This difference persisted when measured at days 11-22 post symptom onset (severe disease 17.30 (6.82-27.32) versus 8.66 (4.25-14.80) in non-severe disease, P=0.016) (Figure 4d, Supplementary table 3).

**Figure 4.**
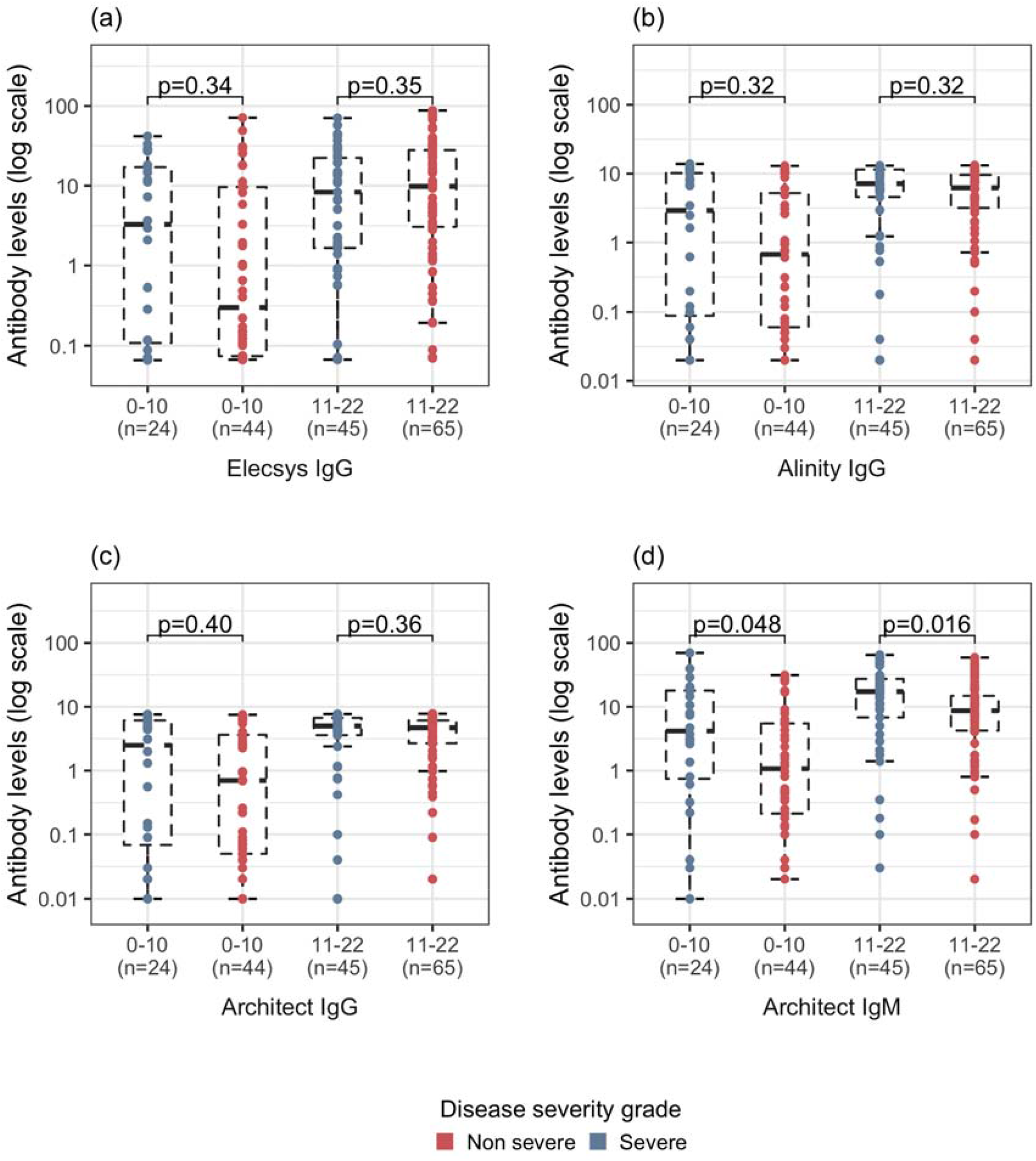
Early antibody levels in subjects who developed severe versus non-severe COVID-19. Log; logarithmic

### Cross-reactivity

Within the SARS-CoV-2 Neg group, 9 (4.02% overall) returned a positive result on the Elecsys assay, of which 8 (3.57% overall) were also positive on both Abbott IgG assays. Detailed clinical review of these 9 subjects revealed that the majority had a clinical presentation suggestive of COVID-19; six presented with an influenza-like illness, two with a systemic inflammatory response, four had history of close contact with a confirmed COVID-19 case and one was diagnosed with a viral myocarditis (Supplementary table 4).

Within the Controls Pre-2020 group, only 1 (0.9%) sample derived from previous flu seasons 2016 to 2019 and 3 samples (1.1%) from historical controls with chronic HIV mono-infection were positive on the Abbott IgG assays. We observed no cross-reactivity with the Elecsys assay for samples from the Controls Pre-2020 group.

## Discussion

This is among the largest and most comprehensive studies analysing the performance and dynamics of both IgG and IgM anti-SARS-CoV-2 assays in well-phenotyped, diverse populations with and without confirmed COVID-19 infection. Our results show a clear delineation between development of IgM and IgG responses, with IgM responses developing earlier after onset of symptoms and predicting development of more severe COVID-19 infection. Furthermore, we demonstrate declines in antibody levels of all assays after a peak that occurs quite early (5-6 weeks) post symptom onset. These data provide important insights into both the clinical utility of serology in managing SARS-CoV-2 infection as well the limited longevity of antibody responses, which may have implications for post-infection immunity to SARS-CoV-2.

Although overall assay sensitivity for all four assays was less than 80%, sensitivity was lower early after symptom onset and increased to levels consistently above 80% after day 11 and maintained beyond day 42 for all but the Abbott IgM assay, which decreased notably after day 42 to 68.5% (Table 3). These data are in keeping with previous studies and metanalyses that demonstrated lower assay sensitivity early after onset of symptoms[7, 11, 13, 16] and other studies that demonstrated high sensitivity in samples taken more than 2 weeks after symptom onset.[9, 15] Data on IgM responses are lacking and limited to relatively small studies,[4, 22] with one study (N=74) showing overall sensitivity of 70% in samples taken at least three weeks post exposure to SARS-CoV-2,[4] similar to that seen in the later time periods of our analysis (>42 days) when the proportion with positive IgM was notably reduced.

To our knowledge this is the first study to map dynamic changes in antibody levels against date of symptom onset within a large, diverse cohort. Consistent across all three IgG-targeted assays, antibody levels peaked just over five weeks after symptom onset and decreased thereafter. The dynamics of IgM titres followed an earlier peak and more rapid subsequent decline, which is biologically plausible. The declines in antibody levels observed across all assays support earlier data from a small cohort that demonstrated loss of both antibody levels and neutralising antibody responses in the early convalescent period[4] and suggest the potential for waning of post-infectious immune responses that may explain the recent increase in reported reinfections with SARS-CoV-2.[23]

In our analysis, significantly higher IgM levels early in infection (before day 10), but not levels of the other antibodies tested, were observed in subjects who developed severe COVID-19. This is in contrast to a previous smaller study that showed higher IgG levels (but not IgM) in subjects with severe compared to asymptomatic SARS-CoV-2 infection, however this previous study did not include as heterogeneous a study population as our analysis. Interestingly, the only other study to report on kinetics of IgM and IgG early into infection (N=23, 11 with moderate and 12 with severe infection) also demonstrated higher early IgM but not IgG levels in more severe disease.[22] These data, confirmed within our large, diverse population suggest a potential role for early measurement of IgM in identifying those presenting with symptoms who are at greater risk of developing severe COVID-19.

Our results add to the body of data showing the high specificity of the serological assays tested. In particular, we showed little cross-reactivity with historical samples from populations with high prevalence of common viral co-infections as well as those taken during previous outbreaks of reported community influenza-like illnesses in Ireland. Of note, when we analysed cases of positive antibody responses in subjects hospitalised but not detected by SARS-CoV-2 PCR we identified a majority that presented with symptoms consistent with COVID-19 where no alternative diagnosis was established. This suggests an additional clinical use for serological testing in aiding clinical diagnosis in these circumstances.

Our study has limitations. Although we measured serological responses, we do not have data on corresponding functional immunity, important when interpreting the clinical relevance of the observed decline in antibody levels. Although we analysed historical samples, we did not have data on confirmed influenza in these subjects nor did we routinely test the SARS-CoV2 Pos and Neg groups for other co-infections.

Despite these limitations, this study, one of the largest and most detailed analyses of the performance and kinetics of anti-SARS-CoV-2 antibody responses, suggests higher, early IgM responses in those who develop more severe COVID-19. The early decline in antibody levels, as early as five weeks post symptom onset, contribute to an increasing concern that post-infectious immunity to SARS-CoV-2 infection may be time limited.

## Data Availability

Aggregate data provided in tables 1 to 3 and supplementary tables 1 to 5. Graphic representation of data provided in figures 1 to 4 and supplementary figures 1 and 2.

## Funding

This work was supported by Science Foundation Ireland [grant number 20/COV/0305] and the European Union’s Horizon 2020 Research and Innovation Programme under the Marie Sklodowska-Curie [grant number 666010 to W.T.]. Abbott Diagnostics provided the reagents for the antibody reactions. The funding sources had no role in the study design, recruitment, data collection or analysis of the study results. Representatives from Abbott Diagnostics were provided an opportunity to review and comment on the manuscript prior to submission.

## Acknowledgements

The authors wish to thank all study participants and their families for their participation and support in the conduct of the All Ireland Infectious Diseases Cohort Study.

## Author contributions

PWG conceived and oversaw the project, provided input into data analysis and interpretation and wrote the manuscript. WT conducted the data analysis, provide interpretation of results and contributed to writing the manuscript. AGL processed and stored samples, coordinated the laboratory analysis and contributed to writing the manuscript. KMcC coordinated site clinical recruitment, clinical data collection and interpretation. GK coordinated site clinical recruitment, clinical data collection, data interpretation and contributed to the writing of the manuscript. PMcG coordinated site clinical recruitment, clinical data collection and interpretation. SG coordinated site clinical recruitment, clinical data collection and interpretation. RI coordinated the laboratory antibody analysis and data interpretation. AGC coordinated site clinical recruitment, clinical data collection and interpretation. ERF coordinated site clinical recruitment, clinical data collection and interpretation. SS coordinated site clinical recruitment, clinical data collection and interpretation. PD conceived the project, oversaw the laboratory antibody analyses and provided input into data analysis, interpretation and manuscript writing.

## Declarations of Interest

Patrick W. G. Mallon has received honoraria and/or travel grants from Gilead Sciences, MSD, Bristol Myers Squibb and ViiV Healthcare.

Willard Tinago – no conflicts declared.

Alejandro Abner Garcia Leon – no conflicts declared.

Kathleen McCann – no conflicts declared.

Grace Kenny – no conflicts declared.

Padraig McGettrick – no conflicts declared.

Sandra Green – no conflicts declared.

Rosanna Inzitiari – no conflicts declared.

Aoife Cotter – no conflicts declared.

Stefano Savinelli – no conflicts declared.

Eoin R Feeney has received honoraria and/or travel grants from Gilead Sciences, Abbvie, MSD, Vidacare and ViiV Healthcare.

Peter Doran – no conflicts declared.

## Study group

### The All Ireland Infectious Diseases Cohort Study

P. Gavin (Children’s Health Ireland). J. Eustace, M. Horgan, C Sadlier (Cork University Hospital). J. Lambert, T. McGinty (Mater Misericordiae University Hospital). J. Low (Our Lady of Lourdes Hospital, Drogheda). B. Whelan (Sligo University Hospital). B. McNicholas (University Hospital Galway). O. Yousif (Wexford General Hospital). G. Courtney (St Luke’s General Hospital Kilkenny). E. DeBarra, C. Kelly (Beaumont University Hospital). T. Bracken (University College Dublin).

## Supplementary tables

**Supplementary table 1.** Comparison of the sensitivity and specificity between the three antibody assays.

| Assay comparison | Sensitivity |  | Specificity |  |
| --- | --- | --- | --- | --- |
|  | %difference (95% CI) | P-value | %difference (95% CI) | P-value |
| Elecsys vs Architect IgG | 0.028 (-0.007; 0.06) | 0.08 | 0.009 (0.0002; 0.018) | 0.046 |
| Elecsys vs Alinity IgG | 0.024 (-0.009; 0.06) | 0.12 | 0.009 (0.0002; 0.018) | 0.046 |
| Architect IgG vs Alinity IgG | -0.003 (-0.012; 0.006) | 0.32 | 0.00 (-0.006; 0.006) | 1.00 |

**Supplementary table 2.** Agreement between assays.

| Antibody immunoassay | Overall percentage agreement | Cohen's kappa |
| --- | --- | --- |
| Elecsys vs Architect-IgG | 96.8 | 0.92 |
| Elecsys vs Alinity-IgG | 96.9 | 0.92 |
| Architect-IgG vs Alinity-IgG | 99.7 | 0.99 |
| *Architect-IgG vs Architect-IgM | 90.5 | 0.79 |

**Supplementary table 3.** Comparison of antibody levels with maximum disease severity attained.

| Assay | Days since symptoms onset |  |  |  |  |  |
| --- | --- | --- | --- | --- | --- | --- |
|  | 0-10 |  |  | 11-21 |  |  |
|  | Non-severe<br>(n=44) | Severe<br>(n=24) | P-value | Non-severe<br>(n=65) | Severe<br>(n=45) | P-value |
| Elecsys IgG | 0.32 (0.07-9.63) | 3.30 (0.11-17.10) | 0.34 | 9.82 (3.06-27.94) | 8.32 (1.66-22.31) | 0.35 |
| Alinity IgG | 0.68 (0.06-5.24) | 2.96 (0.09-10.18) | 0.32 | 6.22 (3.18-9.60) | 7.16 (4.58-11.44) | 0.32 |
| Architect IgG | 0.70 (0.05-3.63) | 2.55 (0.08-6.12) | 0.40 | 4.69 (2.68-6.11) | 5.00 (3.58-6.71) | 0.36 |
| Architect IgM | 1.07 (0.21-5.46) | 4.20 (0.75-17.93) | 0.048 | 8.66 (4.25-14.80) | 17.30 (6.82-27.32) | 0.016 |

**Supplementary Table 4.** Clinical characteristics of subjects in the SARS-CoV-2 Neg group with positive IgG antibody.

| Age range | Gender | Brief description of clinical presentation |
| --- | --- | --- |
| 60-70 | Female | Presented day 8 after symptom onset. Influenza-like illness with fever. NP swab not detected on two occasions. Systemic inflammatory response on bloods. No cause found. |
| 50-60 | Male | Presented with new seizures and cough. No cause found. Previous admission in late 2019 with perihilar parenchymal changes noted on CXR. |
| 70-80 | Male | Presented with clinical diagnosis of pneumonia but not tested for SARS-CoV-2 infection. Serology assessed as part of an outpatient post-COVID ('Long COVID') clinic. |
| 40-50 | Female | Presented 12 days post symptom onset. Influenza-like illness. No cause found. Close contact of a confirmed case. NP not detected on three occasions. |
| 30-40 | Male | Risk through occupational exposure. Close contact of a confirmed case. NP swab negative. |
| 40-50 | Male | Presented with an influenza-like illness and systemic inflammatory response. Multiple NP swabs not detected. High clinical suspicion for COVID-19 infection. |
| 70-80 | Male | Presented with breathlessness, cough and fevers. No cause found but high clinical suspicion. Close contact of a confirmed case (28 days prior to presentation) |
| 30-40 | Female | Presented day 21 after symptom onset with influenza-like illness. NP swab not detected. Diagnosed with viral myocarditis. |
| 50-60 | Male | Risk through occupational exposure. Presented day 14 after symptom onset with cough and breathlessness. WHO Critical Severity grading. NP swab negative on three occasions. High clinical suspicion. |

Age ranges only provided to ensure anonymity. NP; nasopharyngeal. ICU; intensive care unit. CXR; chest x-ray. WHO; World Health Organisation.

**Supplementary table 5.** Specificity of the serological assays in samples taken prior to the 2020 COVID19 pandemic.

| Sera samples cohort | Elecsys | Alinity-IgG | Architect IgG | Architect-IgM |
| --- | --- | --- | --- | --- |
|  | Ab positive<br>n (%) | Ab positive<br>n (%) | Ab positive<br>n (%) | Ab positive<br>n (%) |
| Flu season pre-COVID 19 | 0 (0) | 1 (0.9%) | 1 (0.9%) | 1 (0.9%) |
| HIV | 0 (0) | 3 (1.1%) | 3 (1.1%) | - |
| HIV/HCV | 0 (0) | 0 | 0 | - |
| HIV/HBV | 0 (0) | 0 | 0 | - |
| HBV | 0 (0) | 0 | 0 | - |

## Supplementary figures

**Supplementary figure 1.**
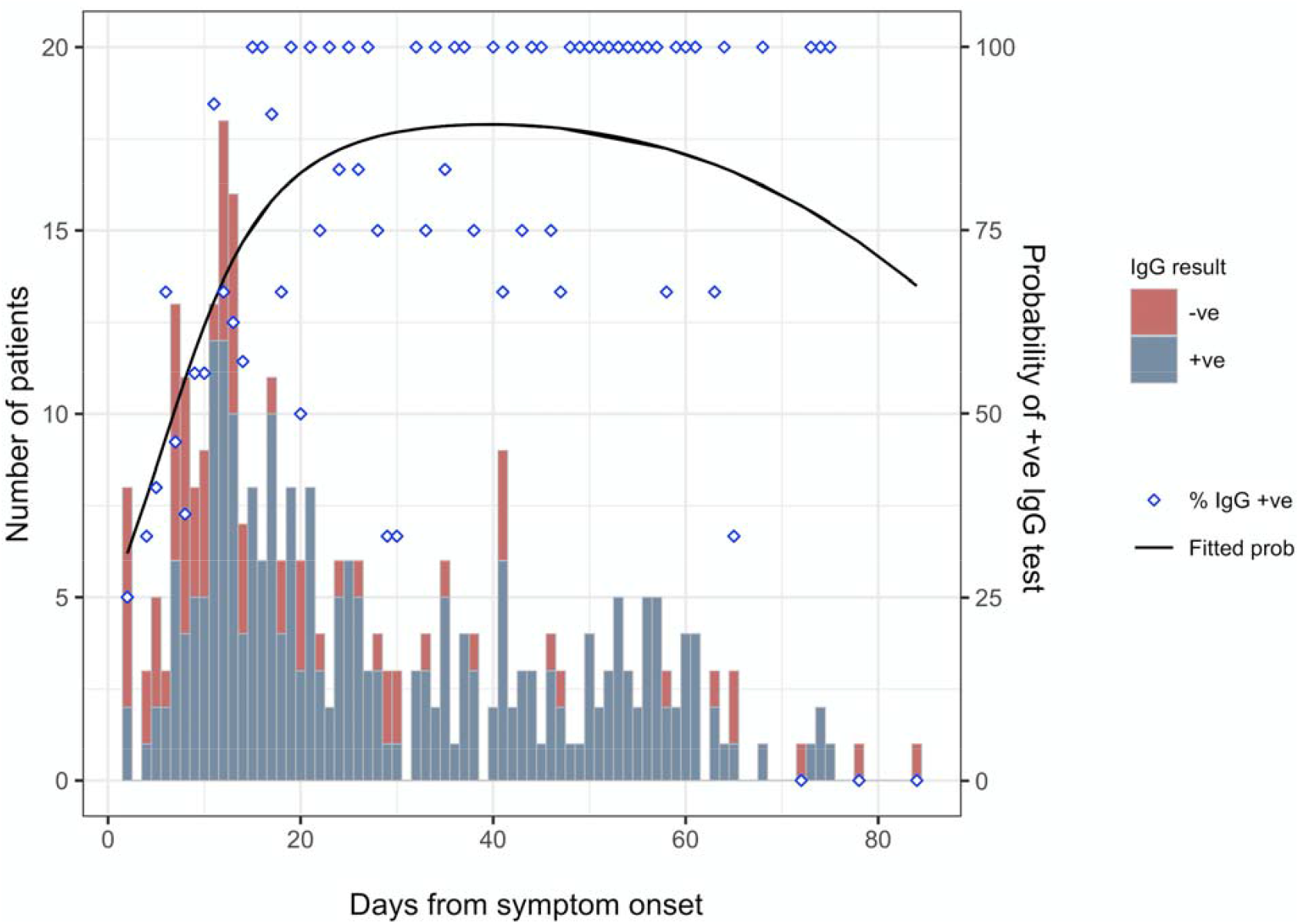
Rates of seropositivity to the Abbott Architect IgG assay and time from symptom onset.

**Supplementary figure 2.**
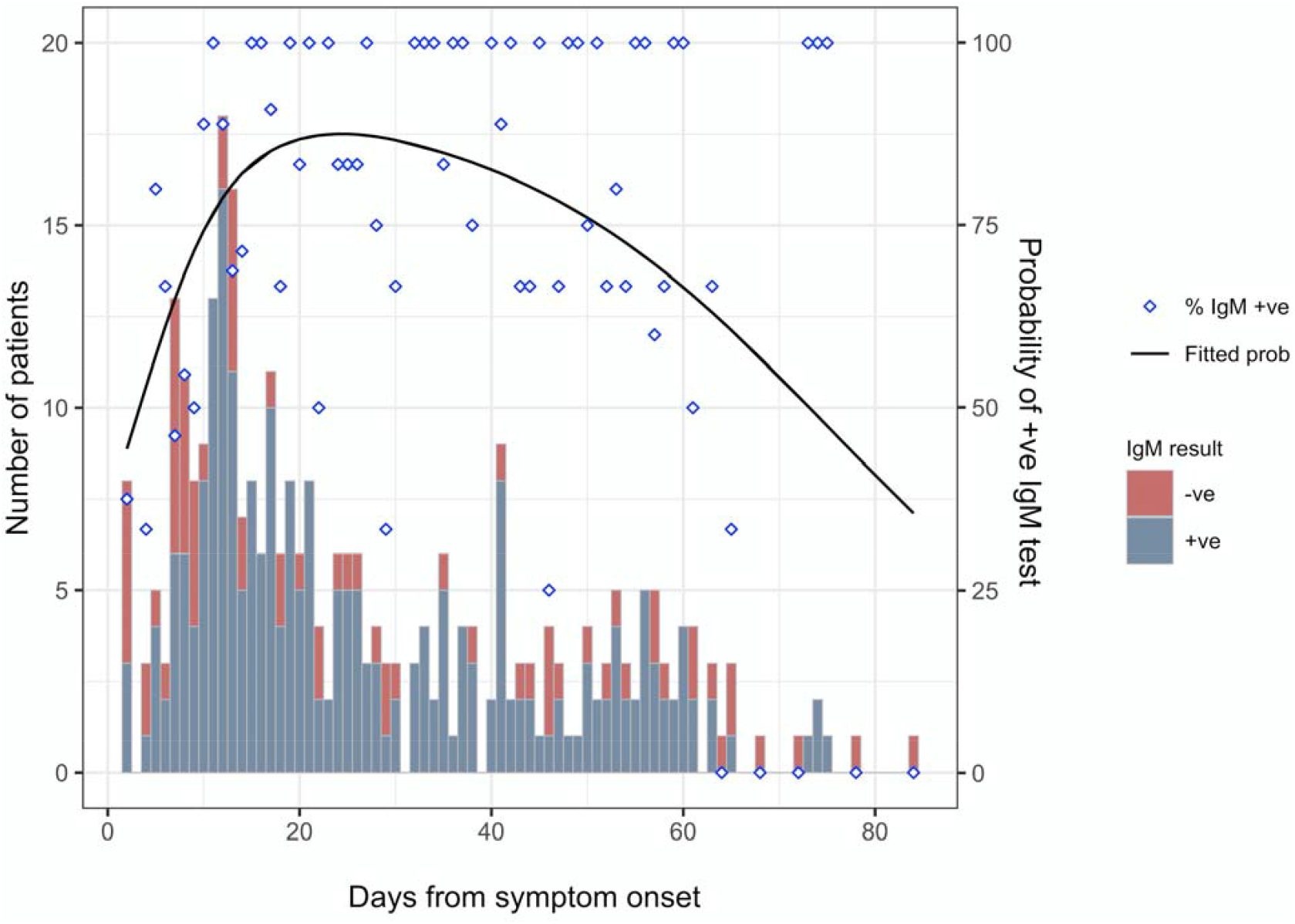
Rates of seropositivity to the Abbott Architect IgM assay and time from symptom onset.

## Notes

### Funding Statement

This work was supported by Science Foundation Ireland [grant number 20/COV/0305] and the European Unions Horizon 2020 Research and Innovation Programme under the Marie Sklodowska Curie [grant number 666010 to W.T.]. Abbott Diagnostics provided the reagents for the antibody reactions. The funding sources had no role in the study design recruitment data collection or analysis of the study results. Representatives from Abbott Diagnostics were provided an opportunity to review and comment on the manuscript prior to submission.

### Author Declarations

1) The Mater Misericordiae University Hospital and Mater Private Hospital Institutional Review Board 2) St. Vincents Healthcare Group Ethics and Medical Research Committee

